# The emergence of SARS-CoV-2 variants of concern is driven by acceleration of the evolutionary rate

**DOI:** 10.1101/2021.08.29.21262799

**Authors:** John H. Tay, Ashleigh F. Porter, Wytamma Wirth, Sebastian Duchene

## Abstract

The ongoing SARS-CoV-2 pandemic has seen an unprecedented amount of rapidly generated genome data. These data have revealed the emergence of lineages with mutations associated to transmissibility and antigenicity, known as variants of concern (VOCs). A striking aspect of VOCs is that many of them involve an unusually large number of defining mutations. Current phylogenetic estimates of the evolutionary rate of SARS-CoV-2 suggest that its genome accrues around 2 mutations per month. However, VOCs can have around 15 defining mutations and it is hypothesised that they emerged over the course of a few months, implying that they must have evolved faster for a period of time. We analysed genome sequence data from the GISAID database to assess whether the emergence of VOCs can be attributed to changes in the evolutionary rate of the virus and whether this pattern can be detected at a phylogenetic level using genome data. We fit a range of molecular clock models and assessed their statistical fit. Our analyses indicate that the emergence of VOCs is driven by an episodic increase in the evolutionary rate of around 4-fold the background phylogenetic rate estimate that may have lasted several weeks or months. These results underscore the importance of monitoring the molecular evolution of the virus as a means of understanding the circumstances under which VOCs may emerge.

## 1 The molecular clock of SARS-CoV-2

Genome sequence data of viruses have been extensively used to track the evolution and spread of these pathogens. The ongoing SARS-CoV-2 pandemic has seen an unprecedented number of genomes generated that have been used to gain rapid insight into epidemiological spread (Dellicour et al., 2021), identify the time of origin (Pekar et al., 2021), and to track mutations of functional importance. Most concerning mutations occur in the spike protein and may increase transmissibility (Kraemer et al., 2021), or disease severity (Harvey et al., 2021), although vaccines are likely still effective against them (Dearlove et al., 2020). Such lineages are known as variants of concern (VOCs) and they are characterised at a genomic level by a number of fixed mutations in the S1 subunit of the spike protein, the most common of which are mutations N501Y and D614G (Eurosurveillance, 2021), with the the latter presenting evidence increased transmissibilty and favoured by selection (Volz et al., 2021). For a lineage to be formally classified as a VOC there must be evidence of an impact in transmissibility, virulence, and/or immunity (Mascola et al., 2021).

SARS-CoV-2 lineages are classified using a dynamic nomenclature system, known as PANGO (Rambaut et al., 2020). Recently the World Health Organisation assigned variants of concern letters of the greek alphabet (Konings et al., 2021). At present the United States CDC recognises four variants of concern; Alpha (PANGO lineage B.1.1.7) first identified in the UK, Beta (PANGO lineage B.1.351) first identified in South Africa, Gamma (PANGO lineage P.1) first identified in Brazil, and Delta (PANGO lineage B.1.617.2) first identified in India (CDC, 2021).

The mechanisms under which VOCs have emerged is not entirely clear, but their defining mutations are well characterised. Variant Alpha has 14 protein-altering mutations and three deletions, with eight of these being in the spike protein. One of the deletions ΔH69/ΔV70 enhances infectivity in vitro and has been detected in immunocompromised patients where immune escape occurred (Kemp et al., 2021, Plante et al., 2021). Variant Beta has nine protein-altering mutations with five altering the receptor binding domain. (Tegally et al., 2021). Variant Gamma has 17 mutations, with 10 found in the spike protein and including N501Y and E484K (Faria et al., 2021). Importantly, Alpha, Beta and Gamma share several important mutations, including N501Y and E404K, which likely enhance affinity to human the ACE2 receptor (Nelson et al., 2021). Variant Delta is characterised by 7 mutations in the spike protein, several of which have been associated with altered immune response and increased viral replication, viral load, likely leading to increased viral fitness (CDC, 2021).

The sheer number of mutations observed in these four VOCs is much higher than what would be expected under phylogenetic estimates of the nucleotide evolutionary rate of SARS-CoV-2, which range from around 7×10^−4^ to 1.1×10^−3^ subs/site/year (Duchene et al., 2020, Ghafari et al., 2021), meaning that only about 2 mutations would accumulate per month along a lineage. In these circumstances, the 14 mutations in Alpha would require a period of at least six months, a time that is inconsistent with its first detection in September 2020, because it would have had to evolve from around March 2020 with most defining mutations undetected for many months.

### 1.1 Bayesian molecular clock models

We investigated whether the emergence of variants of concern is associated with an increase in the evolutionary rate that can be detected using phylogenetic analyses of genome data and in the absence of dense intrahost or transmission chain sampling. To this end, we analysed publicly available nucleotide sequence data from GISAID (Elbe and Buckland-Merrett, 2017, Shu and McCauley, 2017) under a range of molecular clock models that describe the evolutionary rate along branches in phylogenetic trees, shown in the Supplementary material. We consider each model as a hypothesis for which we can assess statistical support using Bayesian model selection techniques. Critically, our analyses do not intend to detect signatures of natural selection, nor to identify genomic regions with higher mutation rates, which have been described elsewhere (Abdool Karim and de Oliveira, 2021, Harvey et al., 2021). Instead, our framework serves to characterise the main patterns of evolutionary rate variation in the genome of the virus that underpin the emergence of VOCs.

The simplest molecular clock model is known as strict molecular clock (SC; Zuckerkandl, 1962, Zuckerkandl and Pauling, 1965) that posits a single evolutionary rate for all branches in a phylogenetic tree, and thus serves as a ‘null’ model. A more complex model is the uncorrelated relaxed clock that assumes that branch rates are independent and identically distributed draws from a statistical distribution (Drummond et al., 2006), for which we considered either a lognormal or a Γ distribution (UCLN and UCG, respectively).

We also considered a range of fixed local clock models (FLC; Yoder and Yang, 2000). These models require an *a priori* definition of a set of ‘background’ branches and a set of branches with different rates, known as ‘foreground’. For example, foreground branches can be defined based on some biological expectation (e.g. Worobey et al., 2014), and thus represent a formal evolutionary hypothesis. The evolutionary rate is constant for a given group of branches, although there exist approaches where branches can be assigned a range of relaxed molecular clocks (Fourment and Darling, 2018). These models differ in their number of parameters and biological assumptions (Supplementary material Table S1; reviewed in Bromham et al., 2018 and Ho and Duchêne, 2014).

We specified six configurations of the FLC model, where the evolutionary rate could vary within VOC clades (FLC clades model in Supplementary material Figure S1) or along the stem (FLC stems+clades), only at stem branches (FLC stems), or where these rates could be shared among all VOCs (FLC shared stems, FLC shared clades and FLC shared clades+stems in Supplementary material Figure S1).

Models in which the rate only changes along the stem branches of VOCs represent a situation where the evolutionary rate may increase for a short period of time before returning to the background rate. In contrast, models where the clade also undergoes a rate change would imply that VOCs have a rate that is statistically different from the background.

An alternative approach to the FLC is the random local clock (RLC; Drummond and Suchard, 2010). The evolutionary rate can change at particular nodes in the tree and the location of such changes and actual rates are inferred. The RLC is a general form of all local clock models, where the simplest form is the strict clock, as a case of no rate changes (Bromham et al., 2018, Ho and Duchêne, 2014).

### 1.2 Bayesian hypothesis testing

We conducted model testing by calculating the log marginal likelihood, a measure of statistical fit, and ranking the models accordingly. The difference in log marginal likelihoods between two models is known as the log Bayes factor (Sinsheimer et al., 1996) and measures the relative support for two models given the data. In general, a log Bayes factor of at least 1.1 is considered as ‘substantial evidence’ in favour of a model, with 2.3 being ‘strong’ and 4.6 ‘decisive’ (Kass and Raftery, 1995). We considered two marginal likelihood estimators, path sampling and stepping-stone sampling Gelman and Meng (1998), Lartillot and Philippe (2006), Xie et al. (2011).

## 2 Results

### 2.1 Model selection

The FLC shared stems model had the highest statistical fit, with a log Bayes factor of over 3 compared to the next best-fitting model (3.85 with path-sampling and 3.97 with stepping-stone sampling; Table 1). The next model with highest log marginal likelihood was the FLC stems. These two models assume that the stem branches of VOC have a rate that differs from the background and they only differ in that the FLC stems model allows each VOC stem branch to have its own rate.

**Table 1:** Model selection results for complete genomes. Estimates of log marginal likelihoods using path sampling and stepping-stone (ps logML and ss logML, respectively). log Bayes factors (BF) are shown for the best-fitting model, relative to all others (larger numbers mean lower statistical fit), and thus they are 0.0 for the top model.

| Model | ps logML | ss logML | ps rank | ss rank | ps BF | ss BF |
| --- | --- | --- | --- | --- | --- | --- |
| FLC shared stems | -55427.65 | -55428.17 | 1 | 1 | 0 | 0 |
| FLC stems | -55431.50 | -55432.14 | 2 | 2 | -3.85 | -3.97 |
| UCG | -55433.64 | -55434.26 | 3 | 3 | -6.00 | -6.01 |
| UCLN | -55434.32 | -55434.69 | 4 | 4 | -6.67 | -6.52 |
| FLC shared clades+stems | -55443.30 | -55443.50 | 5 | 5 | -15.64 | -15.34 |
| SC | -55443.53 | -55444.21 | 6 | 6 | -15.88 | -16.04 |
| FLC shared clades | -55449.89 | -55450.52 | 7 | 7 | -22.23 | -22.35 |
| FLC clades+stems | -55453.91 | -55454.58 | 8 | 8 | -26.25 | -26.41 |
| FLC | -55461.87 | -55462.54 | 9 | 9 | -34.21 | -34.38 |

The uncorrelated relaxed clocks had very similar performance, but ranked well below the best model, with a log Bayes factor of at least -6 relative to the FLC shared stems model 1. The log Bayes factors for the remaining models were at least -15, implying ‘strong’ evidence against them, relative to the FLC shared stems.

Interestingly, FLC models where VOC clades were defined as foreground had decisively lower statistical performance than those where only stem branches were labelled as foreground (Table 1). In fact, even the SC model, which is generally considered unrealistic for empirical data, had a log Bayes factor of at least 4 with respect to FLC shared clades and the FLC clades+stems Table 1.

### 2.2 Rates of evolution of variants of concern

The FLC shared stems model had a mean background evolutionary rate of 0.58×10^−3^ subs/site/year (95% CI: 0.51 - 0.65×10^−3^), while that for the VOC stems was 2.45×10^−3^ subs/site/year (95% CI: 1.15 - 4.72×10^−3^). As such, the VOC stems rate was around 4 fold higher than the background (mean 4.25, 95% CI: 2.61 - 8.19) (Fig 1).

**Figure 1:**
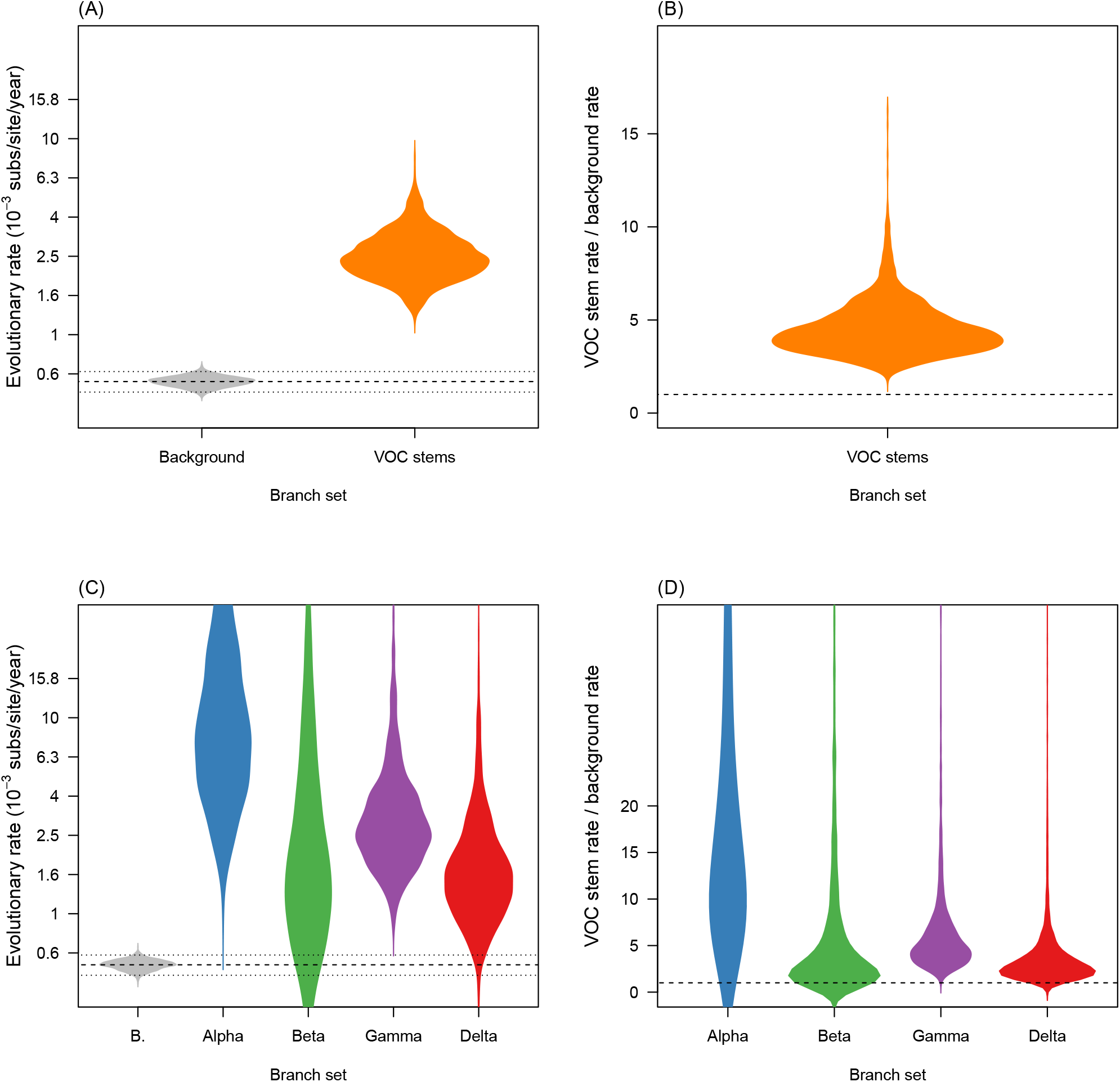
Violin plots for posterior statistics of fixed local clock models (FLC). (A) is for a FLC where the stem branches of VOCs share an evolutionary rate that is different to that of the background (model ‘FLC shared stems’ in Supplementary material Table S1 and Fig S1. The evolutionary rate for variants of concern (VOC) stem branches is shown in orange and the background in grey. The dashed line represents the mean background rate and the dotted lines are the 95% credible interval. (B) is the ratio of the evolutionary rate for VOC stem branches and the background under the same model and the dashed line represents a value of 1.0 where the background and VOC stem rate would be the same. (C) and (D) show the corresponding statistics for the FLC stems model, where the stem branch of every VOC has a different rate. Abbreviation ‘B.’ stands for background.

Although the FLC stems model that assigned each VOC stem branch a different rate had very high uncertainty, it also suggested much higher rates for these branches. The mean background rate under this model was 0.55×10^−3^ subs/site/year (95% CI: 0.49 - 0.62×10^−3^). The corresponding values for VOC were 8.47×10^−3^ subs/site/year (95% CI: 1.93 - 82.37×10^−3^) for Alpha, 1.71×10^−3^ (95% CI: 0.34 - 33.20×10^−3^) for Beta, 2.76×10^−3^ (95% CI: 1.21 - 13.23×10^−3^) for Gamma, and 1.54×10^−3^ (95% CI: 0.62 - 7.35×10^−3^) for Delta.

Clearly, these estimates were several fold higher than that of the background branches, and in spite of their high uncertainty least 0.90 of the posterior density was above the mean background rate (Fig 1).

The coefficient of rate variation for both relaxed clock models, UCG and UCLN, was indicative of departure from clocklike evolution in the data. To investigate whether VOC stem branch rates differed from the rest, we extracted individual branch rates and compared the VOC stem branch rates to the mean of all other branches. We found evidence that VOC stem branch rates were higher than the mean of other branches, with higher means values, but very high uncertainty and 95% credible intervals that overlapped with the mean of other branches (Fig 2).

**Figure 2:**
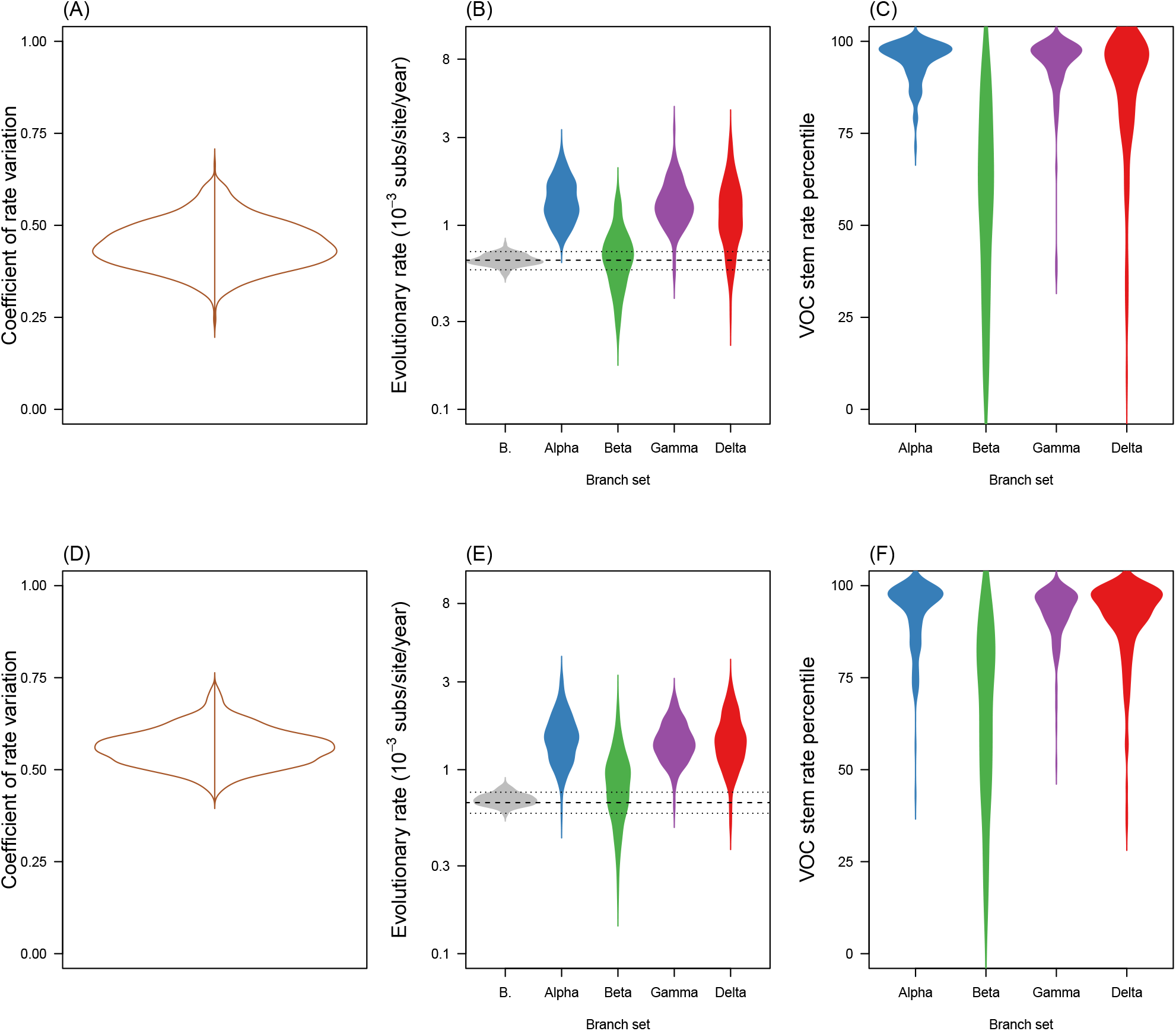
Violin plots of posterior statistics for the uncorrelated relaxed clocks with lognormal (UCLN) and gamma (UCG) distributions (see Supplementary material). The top row, (A) through (C), is for the UCLN and the bottom row, (D) through (F), is for the UCG. (A) and (D) show the coefficient of rate variation, which is the standard deviation of branch rates divided by the mean rate, and indicates clocklike behaviour when it is abutting zero (Drummond et al., 2006, Ho et al., 2015). In (B) and (E) the evolutionary rate is shown for the stem branches of variants of concern (VOC) and for the mean of background branches (i.e. those that are not the stems of VOCs), abbreviated as ‘B.’. The dashed line denotes the mean background rate, while the dotted lines represent the upper and lower 95% credible interval. (C) and (F) shows the percentile in which stem branches for VOCs fall with respect to other branches. Note that the densities have been smoothed, but the maximum values are 100.

The mean evolutionary rate of branches other than the VOC stems was 0.65×10^−3^ subs/site/year (95% CI: 0.58 - 0.77×10^−3^) in the UCLN and 0.69 ×10^−3^ subs/site/year (95% CI: 0.60 - 0.80×10^−3^) for the UCG. In contrast, the VOC stem mean evolutionary rates for the UCLN were: 1.29×10^−3^ subs/site/year (95% credible interval, CI: 0.76 - 2.56×10^−3^) for Alpha, 0.64×10^−3^ (95% CI: 0.32 - 1.57×10^−3^) for Beta, 1.29×10^−3^ (0.82 - 2.40×10^−3^) for Gamma, and 1.06×10^−3^ (95% CI: 0.50 - 2.38×10^−3^) for Delta, and with comparable values for the UCG. The percentile where VOC stems rates fell with respect to other branches also supported the finding that their rates were particularly high in most cases. In the UCLN, for Alpha 0.96 of posterior density had the stem rate in the top 75% of fastest evolving branches, with the corresponding numbers for the other VOCs being 0.25, 0.98, and 0.81 Beta, Gamma, and Delta, respectively, and with comparable values in the UCG (0.92, 0.45, 0.96, and 0.91).

The RLC model produced less clear results than the other molecular clock models. The maximum *a posteriori* number of rate changes was 4, with the 95% CI ranging between 2 and 5. However, the posterior probability of rate changes in VOC stem branches or clades was 0.0. Instead, rate changes were not consistently found on particular branches. It is conceivable that this model poses a heavy penalty on rate changes. In particular, there is a very large number of local clock configurations in these data, which may be impossible to visit under this model and may result in low statistical power to assess support for our hypotheses here. This model, however, had an evolutionary rate estimate that was comparable to that of other models (mean 0.60×10^−3^ subs/site/year; 95% CI: 0.49 - 0.72×10^−3^).

### 2.3 Emergence time and expected genome substitutions

We estimated the duration of time along VOC stem branches and the inferred total number of nucleotide substitutions along the complete genome. We focus on the best fitting model (FLC shared stems), with similar results for the second best model (FLC stems). The duration of time along these branches represents the time required before VOCs started to diversify, but it is important to note that they are contingent on sampling bias, and could therefore be shorter than estimated here. Under the FLC shared stems model, the stem branch leading up to VOs were; 14 weeks (95% CI:6 - 24) for Alpha, 4 (95% CI: 2 - 8) for Beta, 17 (95% CI: 8 - 28) for Gamma, and 6 (3 - 11) for Delta (Supplementary material Fig S2).

The expected number of substitutions along the complete genome were; 21 (95% CI: 14 - 32) for Alpha, 6 (95% CI:3 - 11) for Beta, 26 (95% CI: 18 - 35) for Gamma, and 9 (95% CI: 6 - 16) for Delta. Although, these numbers are loosely associated with the defining mutations, they are not directly comparable because they involve substitutions along the entire genome and they correspond to the inference from a standard phylogenetic substitution model (the GTR+Γ in this case).

## 3 Discussion

Our mean rate estimates over all lineages are somewhat lower than earlier estimates (Duchene et al., 2020), which is consistent with the notion that the virus has had time to evolve and remove transient deleterious mutations since its emergence (Ghafari et al., 2021). However, the molecular evolutionary rate of SARS-CoV-2 displays substantial variation among lineages, a pattern that has been apparent since early phylogenetic analyses of the virus (Duchene et al., 2020). Evolutionary rate variation is sometimes stochastic in nature and pinpointing its causes is often difficult in empirical data.

Our explicit hypothesis testing framework suggest that the emergence of VOCs explains much of the evolutionary rate variation in the virus. This model testing framework has been previously used to understand viral evolution among host species in influenza (Worobey et al., 2014), and the host range SARS-CoV-2 and closely related viruses (MacLean et al., 2021). We suggest that model testing may be preferable to using highly parametric models, such as relaxed molecular clock models for this purpose, because they tend to have very high variance in parameters of interest, such as evolutionary rates of particular branches. Recent advances in random local clock models may provide increased sensitivity (Fisher et al., 2021) of this family of models.

We find compelling evidence that episodic, instead of long-term, increases in the evolutionary rate un-derpin the emergence of VOCs, a process that is probably driven the action of natural selection. All models where VOC clades were assigned a different rate to the background had poor statistical fit, even when compared to the SC ‘null’ model, providing further support for such rate increases to occur over a short period of time. The increase in evolutionary rate required to give rise to the four VOCs examined was estimated to be around 4-fold compared to the background, although such estimates may carry high uncertainty when estimated for individual stem branches. Under these circumstances the number of mutations required to give rise to a VOC, such as Alpha, would have accumulated in about three months, with some variants requiring less a few weeks, such as Beta and Delta. These timescales appear plausible in chronic infections of SARS-CoV-2 (Harvey et al., 2021, Kemp et al., 2021), but other circumstances are also likely, such as when transmission low and selection favours mutations that increase transmissibility between hosts.

Our genomic analyses demonstrate that signatures of increased evolutionary rates are detectable using phylogenetic methods and genome surveillance data, but the precise mechanism (ecological or intrahost) of how VOCs have emerged is still unclear. To elucidate these processes will require dense sampling between transmission chains, specifically in settings where transmission is unlikely and intra-host sequence data is available. An other important area that is currently under intense investigation is how natural selection shapes the emergence and persistence of VOCs (Tegally et al., 2021). Such studies may benefit from using explicit codon evolution models and careful partitioning among genes to model mutational heterogeneity. We recommend that further research focuses on early detection and understanding of the circumstances under which viral lineages with epidemiological impacts, such as VOCs, emerge.

## 4 Materials and methods

### 4.1 Data set construction

We downloaded 100 randomly selected sequences from the latest global NextStrain SARS-CoV-2 build (Hadfield et al., 2018), from the GISAID database (Elbe and Buckland-Merrett, 2017, Shu and McCauley, 2017). This set of sequences did not include any of those belonging to the four VOCs (Alpha, Beta, Gamma, or Delta) and we also excluded samples drawn from non human hosts. We downloaded 20 randomly selected sequences from the four VOCs to generate a data set of 180 genomes, which we aligned using MAFFT (Katoh and Standley, 2013). Importantly, we ensured that the sequences consisted of complete genomes, with no stretches of more than 10 Ns and excluding those with low coverage (see Supplementary material). To verify that samples classified as VOCs were correctly assigned as such, we estimated a phylogenetic tree using maximum likelihood as implemented in IQ-TREE2 (Minh et al., 2020), using the GTR+Γ substitution model and with approximate Bayes branch support (Anisimova et al., 2011). We ensured that all VOC samples that were monophyletic with other VOC samples with an approximate Bayes support *<*0.95.

### 4.2 Bayesian phylogenetic analyses

Our Bayesian analyses require specifying a substitution model, a tree, prior, priors for all parameters in BEAST 1.10 (Suchard et al., 2018). We chose the GTR+Γ_4_ substitution model and a coalescent exponential tree prior. Although the tree prior is not necessarily realistic here, it is expected to have little impact in molecular clock estimates Ritchie et al. (2017). It can accommodate changes in population size via the exponential growth function and it is fully parametric, such that setting proper priors for all parameters is possible. To calibrate the molecular clock we specified the sequence sampling times. The FLC models require constraining monophyly in VOCs, which we also did for other clock models to ensure that the prior on tree topology was the same.

We used the default priors for the substitution model. The coalescent exponential tree prior has two parameters, the scaled population size, Φ, and the growth rate *r*. The scaled population size is proportional to the number of infected individuals at present divided by the twice the coalescent rate, *λ*, (i.e. 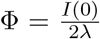) and the growth rate is inversely proportional to the doubling time by a factor of log(2) 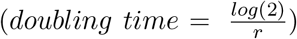 (Boskova et al., 2014, Volz et al., 2009). We used priors with relatively low information content for these two parameters. For Φ we used an exponential distribution with mean 10^5^, while for *r* we used a Laplace distribution with location 0 and scale 100. For all molecular clock rates we used a continuous-time Markov chain reference prior (Ferreira and Suchard, 2008). The UCLN and UCG models have an additional parameter; the standard deviation of the lognormal distribution, and the shape of the Γ distribution. For these parameters we specified an exponential prior with mean 0.33. We ran our analyses for using a Markov chain Monte Carlo of length 5×10^7^, sampling every 5×10^3^ and discarding 10% of the chain as burn-in. We repeated the analyses once to verify convergence of independent chains and we ensured that the effective sample size of all parameters was at least 200.

### 4.3 Marginal likelihood estimation

We used two techniques to infer the log marginal likelihood; path-sampling and stepping-stone (Gelman and Meng, 1998, Lartillot and Philippe, 2006, Xie et al., 2011), which have been found to have high performance in differentiating models in phylogenetics (Baele et al., 2012, 2013, Fourment et al., 2020), reviewed by Baele and Lemey (2014), Oaks et al. (2019). We chose these estimators over the more recently developed and highly accurate generalised stepping-stone because it requires a working genealogical distribution (Baele et al., 2016), which is not trivial here due to the monophyletic constraints. Our estimation setup had 200 path steps distributed according to quantiles from a *β* distribution with parameter *α*=0.3, with each of the resulting 201 power posterior inferences running for 10^6^ iterations. We repeated these analyses three times to assess their variance. Our model testing approach considered the UCLN, SC, and all FLC models in Table 1 and Supplementary material. We did not calculate log marginal likelihoods for the RLC because this is a model averaging method, where the number of parameters is less tractable than in other models. As a result it is difficult to conceive proper priors for all parameters, which is a fundamental aspect of Bayesian model selection.

## Supporting information

Acknowledgements table

## Data Availability

All data used in this study are available through the GISDAID database, with accession numbers reported in supplementary material.

## 5 Acknowledgements

This work was supported by the Australian Research Council (DE190100805) and the Medical Research Future Fund (MRF9200006). This research was undertaken using the LIEF HPC-GPGPU Facility hosted at the University of Melbourne. This Facility was established with the assistance of LIEF Grant LE170100200. We acknowledge efforts by originating and submitting laboratories for the sequence data in GISAID EpiCoV on which our analyses are based. We are also grateful to Prof. Edward Holmes for useful suggestions and comments on ideas developed in this study.

