## Supplementary material for "The emergence of SARS-CoV-2 variants of concern is driven by acceleration of the evolutionary rate"

### 1 Supplementary material

GISAID acknowledgements table

Table S1: Molecular clock model configurations and parameterisation. The term 'clades' correspond to monophyletic groups of either of the four variants of concern (VOC) in the data set (Alpha, Beta, Gamma, Delta) and the stems are the branches leading up to them. Note that 'clock rate' refers to the global clock rates of the strict clock and 'b. rate' is for the background rate.

| **Model family** | **Model abbreviation** | **Parameters** | **Number of parameters** |
|---|---|---|---|
| Strict clock | SC | clock rate | 1 |
| Relaxed | UCLN | Num. branches, mean, s.d. | 358+2 |
| Relaxed | UGM | Num. branches, mean, shape | 358+2 |
| Random local clock | RLC* | b. rate, num. rate changes | variable |
| Fixed local clock | FLC clades | b. rate, num. clades | 1+4 |
| Fixed local clock | FLC stems+clades | b. rate, num. clades & stems | 1+4 |
| Fixed local clock | FLC stems only | b. rate, num. stems | 1+4 |
| Fixed local clock | FLC shared stem | b. rate, stem rate | 1+1 |
| Fixed local clock | FLC shared clade | b. rate, clade rate | 1+1 |
| Fixed local clock | FLC shared clades+stems | b. rate, clade & stem rate | 1+1 |

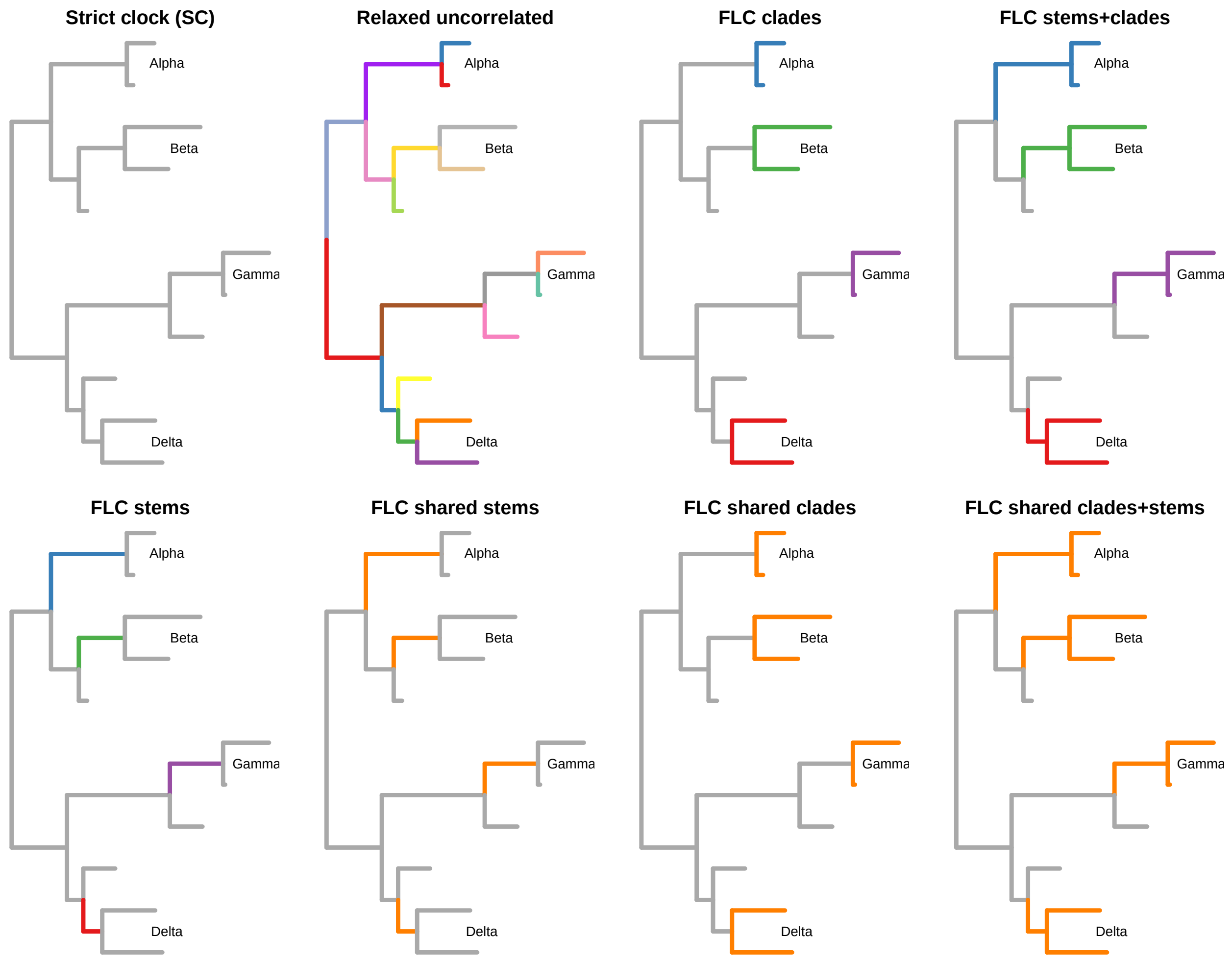


Figure S1: Illustration of molecular clock models considered in hypothetical trees with the four variants of concern (VOCs) and background genetic diversity. SC stands for strict clock and FLC for fixed local clock. Branches are coloured according to the rate assigned in each case, with grey corresponding to the 'background' branches and those in colours being the 'foreground' branches. Model names match those of Supplementary material Table 1.

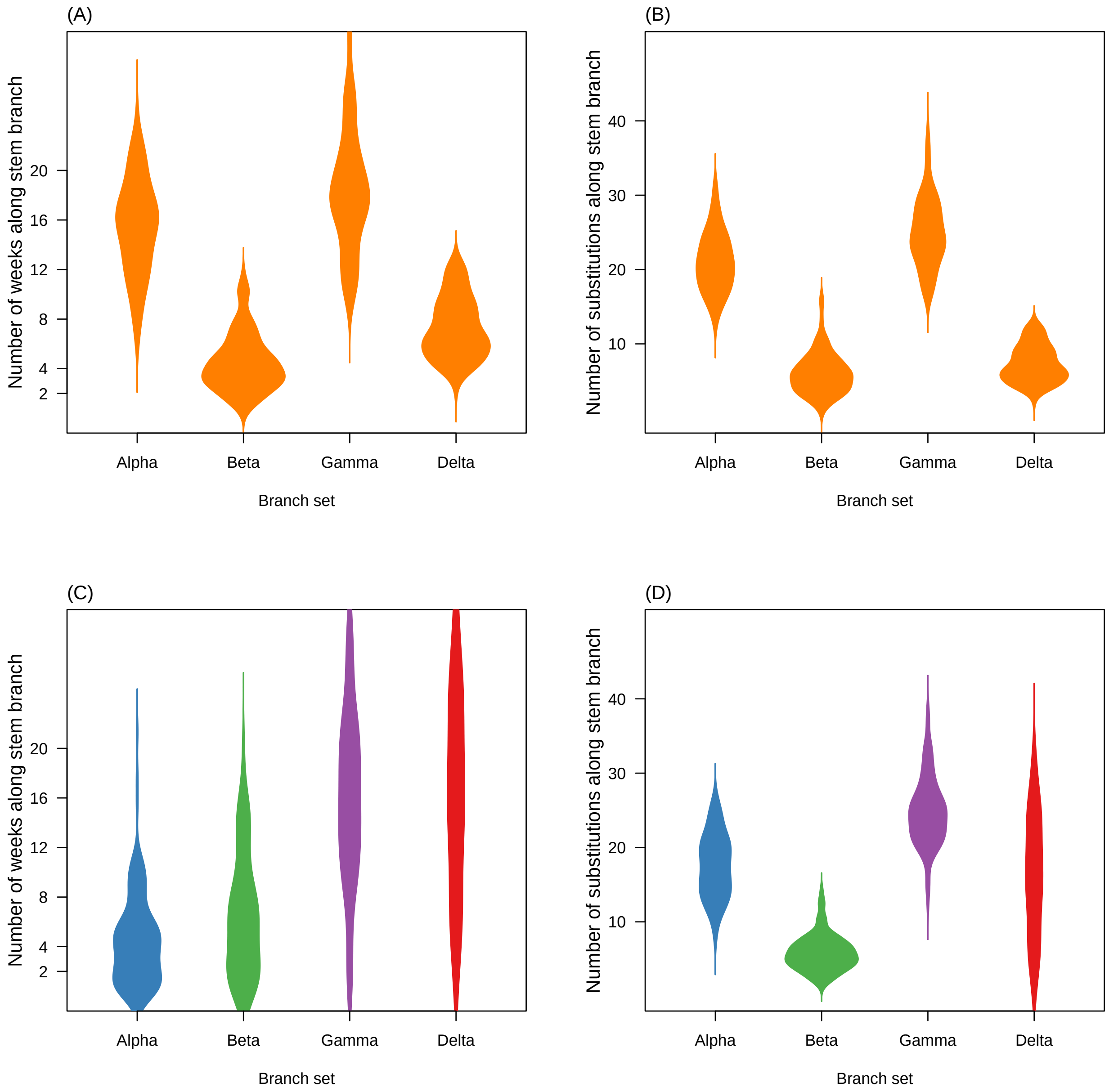


Figure S2: Violin plots of the number of weeks and expected number of substitutions along VOC stem branches for the FLC shared stems model (panels (A) and (B)) and FLC stems (panels (C) and (D)).
